# Can we distinguish the roles of demographic and temporal changes in the incidence and prevalence of musculoskeletal disorders? A systematic search and review

**DOI:** 10.1101/2021.09.20.21263840

**Authors:** Hanifa Bouziri, Alexis Descatha, Yves Roquelaure, William Dab, Kévin Jean

## Abstract

**Background:** Musculoskeletal disorders (MSDs) represent a major public health issue, affecting more than 40 million European workers in 2017. The overall ageing of the working population is expected to increase the burden of disease, but temporal changes in exposures or diagnosis may also drive global trends in MSDs. We, therefore, sought to review and summarize evidence describing the role of demographic and temporal changes in the occurrence of MSDs.

**Methods:** We conducted a systematic search and review of articles reporting temporal trends in MSDs in the general working-age population. Only articles controlling for age in the analysis were included. The risk of bias was assessed. The main indicators extracted were age-controlled time trends in MSD incidence or prevalence.

**Results:** Among 2,680 articles, 16 fulfilled the inclusion criteria, representing 23 results according to the indicators extracted. No study was found with a high risk of bias. Twelve results reported time trends in prevalence and 11 in incidence. After controlling for age, the reported temporal trends varied, mostly between non-monotonic changes (n=12/23) and increases (n=10/23); one article also highlighted an increase among women and non-monotonic changes among men (n = 1/23). Several factors other than ageing were suggested to explain temporal trends in MSDs, main trends in obesity, changing occupational exposures, and cultural factors regarding pain tolerance.

**Conclusion:** This review shows that different kinds of factors in addition to ageing may contribute to varying or increasing trends in MSDs. This review also highlighted the scarcity of evidence regarding time trends in the burden of MSDs and their underlying causes.

## INTRODUCTION

Musculoskeletal disorders (MSDs) affected more than 40 million workers in Europe in 2017 and were the leading contributors to disability worldwide in 2019 (1, 2). These conditions refer to a group of painful disorders of muscles, tendons, and nerves. The causes of MSDs are multifactorial and notably can be induced by occupational, biomechanical and psychosocial risk factors (3). Since the 1970s, there have been many changes in working conditions, catalyzed most notably by increased digitization across a range of professions, and widespread reinforcement of preventive actions has redistributed the risk factors of MSDs (4-6). There has also been an increase in employment in the service sector, which contributes to changes in the patterns of exposure to hazards at work (1). The combined effects of these occupational changes on the temporal evolution of MSDs are thus challenging to assess. In addition, the ageing of the workforce could have implications for the increasing risk of chronic diseases like MSDs (6, 7). In particular, the rising average age of workers in many high-income countries may increase the risk of MSDs in the absence of preventive action. This owes in part to degenerative phenomena linked to the ageing process itself, which induces a reduction in biomechanical tolerance to repetitive and/or prolonged loading, and in part to prolonged exposure to residual biomechanical stresses and psychosocial risks accumulated during increasingly long careers (3).

Exposures to leading occupational risk factors of MSDs, such as biomechanical, organizational, and psychosocial factors, have evolved heterogeneously over time (8-11). Consequently, successive cohorts of workers have not been exposed to MSD risk factors with the same intensity and frequency throughout their lives. Moreover, risk factors of MSDs and their changes over time have mainly been studied individually or by family (e.g. biomechanical, organizational, psychosocial), but a global view of their simultaneous change over time is still lacking. Disentangling the respective roles of age and temporal evolution in exposure to risk factors on the occurrence of MSDs is thus needed to understand current trends and design adapted prevention policies (3-14). Furthermore, understanding the evolution of exposures over time while accounting for age would allow for a more accurate prediction of future trends in MSDs and help to prevent and control their occurrence (15, 16).

The objective of this study is to collate and review the existing evidence on the respective roles of demographic and temporal changes in the occurrence of MSDs. We used the systematic search and review methodology as previously described by Grant and Booth (17). This type of review consists of combining a systematic search method with a critical review analysis and is used to answer broad questions while often incorporating multiple study designs.

## METHODS

### Search strategy

The study protocol was registered in PROSPERO (CRD42020221499) (18). This protocol is consistent with the Preferred Reporting Items for Systematic Review and Meta-analysis (PRISMA) guidelines (19, 21). A detailed PRISMA 2020 checklist is provided in supplementary material 1. Any modification of the methods stated in the original protocol was registered in PROSPERO (see the reference mentioned before).

### Literature search

We searched four different electronic bibliographic databases for studies published between 1990 and 2020: Medline, ScienceDirect, Wiley, and Web of Science. The last source searched or consulted was checked in November 2020. Details of the search strategy used for each database are provided in supplementary material S2, including the algorithms of keywords used, the number of results and the articles preselected for screening.

### Inclusion and exclusion criteria

For the article identification step, when the databases allowed, we automatically excluded results related to topics not relevant for our search, such as studies involving animals, molecular biology, immunology studies or clinical case reports.

Then, the first round of selective screening was carried out based on titles and abstracts (step 1). Only original articles were included; conference reports, literature reviews, and editorials were excluded. At this stage, only articles that reported MSD or MSD proxy outcomes while mentioning the notion of temporal trends were included.

In the full-text assessment (step 2), articles defining MSD as a group or set of diseases localized at or around the joints (wrists, elbows, shoulders, spine, or knees) were selected. The pathologies considered here concerned the muscles, tendons and tendon sheaths, nerves, bursae, joints, ligaments, at the periphery of the joints of the upper limbs, the spine, and the lower limbs. We excluded MSDs defined as a joint manifestation of organic diseases (e.g. psoriasis, lupus, gout, etc.) or as the joint location of systemic inflammatory origins (e.g. secondary osteoarthritis). At this step, only articles reporting temporal trends in incidence and/or prevalence in MSD while controlling for age were selected. We included studies conducted among the working-age population. Studies of people under 18 and unpaid domestic workers were excluded. The prevalence or incidence of MSDs over time that only addresses the average over a single period were also excluded.

### Screening

The Covidence Systematic Review software allowed the selection of studies, their download, and the removal of duplicates (22). Both steps 1 and 2 were performed by two independent authors to assess the eligibility of studies identified in the databases. Any conflict in article screening or full-text assessment was resolved by a third senior researcher.

### Data extraction

All articles included were read for the identification and extraction of the following characteristics: geographic location, population studied, study design and recruitment criteria, start and end date of follow-up, MSD sites (superior limbs, inferior limbs & back, or not specified), criteria used for MSD definition (either based on pain or disability), and the method used for MSD diagnosis.

### Assessing risk of bias and quality of evidence

To assess the risk of bias across included studies, we used the RoB-SPEO (23) and the Navigator guide tool, which we adapted for our study (see supplementary material 4 for methodological details) (24, 25). The biases we assessed were selection bias, potential biases linked to misclassification of MSDs, biases due to incorrectly taking confounding factors into account, and bias due to potential conflict of interest. Each article has been classified according to its level of bias (low, probably low, probably high, high). We also assessed the quality of the statistical trends tested by using the following classification: satisfactory quality, probably satisfactory quality, probably unsatisfactory quality, unsatisfactory quality. Further details on the criteria and classifications used for the risk of bias and quality of evidence assessment are available in supplementary material S4.

### Analysis of the temporal trends of the occurrence of MSDs

The following data were extracted from each article: the raw temporal trends of MSD prevalence and/or incidence (if reported), information concerning methods used to control for age, and MSD prevalence and/or incidence over time after controlling for age. For each article, the temporal trends in MSD prevalence and/or incidence were analyzed according to the location and severity of MSDs. If an article investigated multiple types of MSDs and/or addressed both temporal trends in the prevalence and incidence of MSDs, we considered these results independently; therefore, the total number of results could possibly be higher than the number of studies included. We also distinguished two groups of results based on the MSD sites and criteria used for MSD definition, either based on pain or repercussion on work and/or social life (hereafter called disability). The precise definition of MSD used in each article is provided in supplementary material S3. Temporal changes in MSD prevalence/incidence were summarized according to whether they decreased, varied non-monotonically, or increased.

### Synthesized evidence

For articles reporting both raw and age-adjusted MSD time trends, we compared findings to discern differences between them, and therefore to assess whether it would be possible to dissociate age from time in the occurrence of MSDs over time. When mentioned, we summarized the interpretations and hypotheses proposed to explain observed temporal trends in MSD prevalence/incidence.

## RESULTS

### Studies selected

A total of 2,680 study records were identified through our systematic search, of which 1,977 were excluded, 335 were duplicates, and 1,642 were deemed irrelevant as per automatic categorization tools provided by some databases (**Figure 1**). A further 658 records were excluded after the title and abstract screening because they did not present original results and/or did not report results based on MSDs and/or did not address temporal trends in their occurrence. Of the 45 full-text articles assessed for eligibility, 29 were classified as non-eligible, 13 of which because they did not control for age is reported indicators. A total of 16 studies fulfilled all eligibility criteria and were thus included in the review.

**Figure 1:**
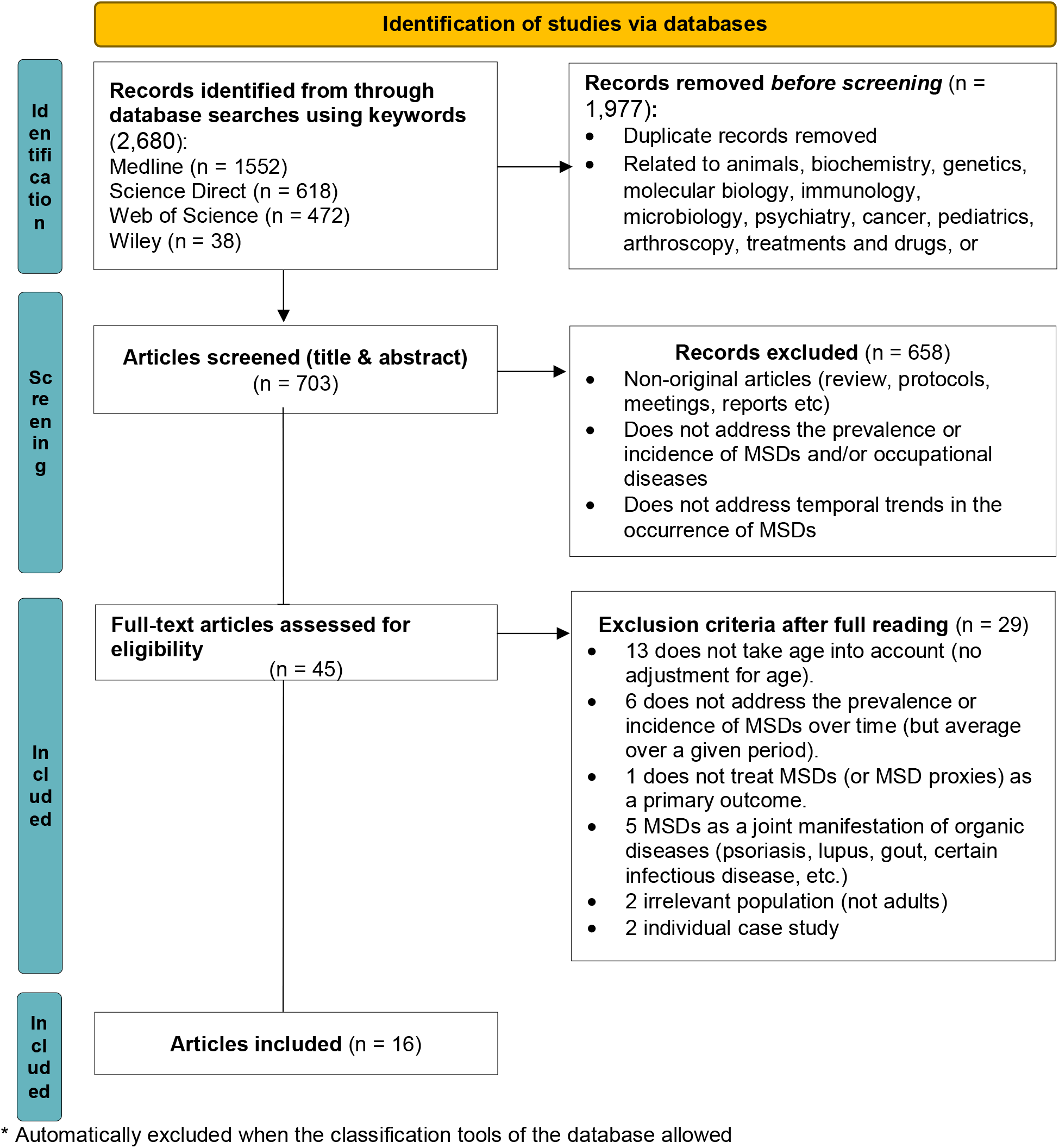
Flow chart diagram of study selection using PRISMA Flow Diagram recommendations. *Inspired by* Page MJ, McKenzie JE, Bossuyt PM, Boutron I, Hoffmann TC, Mulrow CD, et al. The PRISMA 2020 statement: an updated guideline for reporting systematic reviews. BMJ 2021;372:n71. doi: 10.1136/bmj.n71. For more information, visit: http://www.prisma-statement.org/

### General study characteristics

The 16 articles included in the present review were published between 2003 and 2020 (**Table 1**), among which 12 were published after 2010. Overall, the studies were conducted in 3 geographic areas: 12 in Europe (among which 5 were in Scandinavian countries), 3 in the USA, and 1 in Australia. The duration of the study period ranged from 10 to 55 years across studies. Eight studies relied on sampled populations (3 cohort designs, 5 repeated cross-sectional studies) representing a total of 1,387,930 individual working-age adults. Among these, 2 studies focused on the male population only. The other 8 studies relied on a time-series design based on surveillance data collected within 5 countries and 1 subnational administrative area.

**Table 1:**
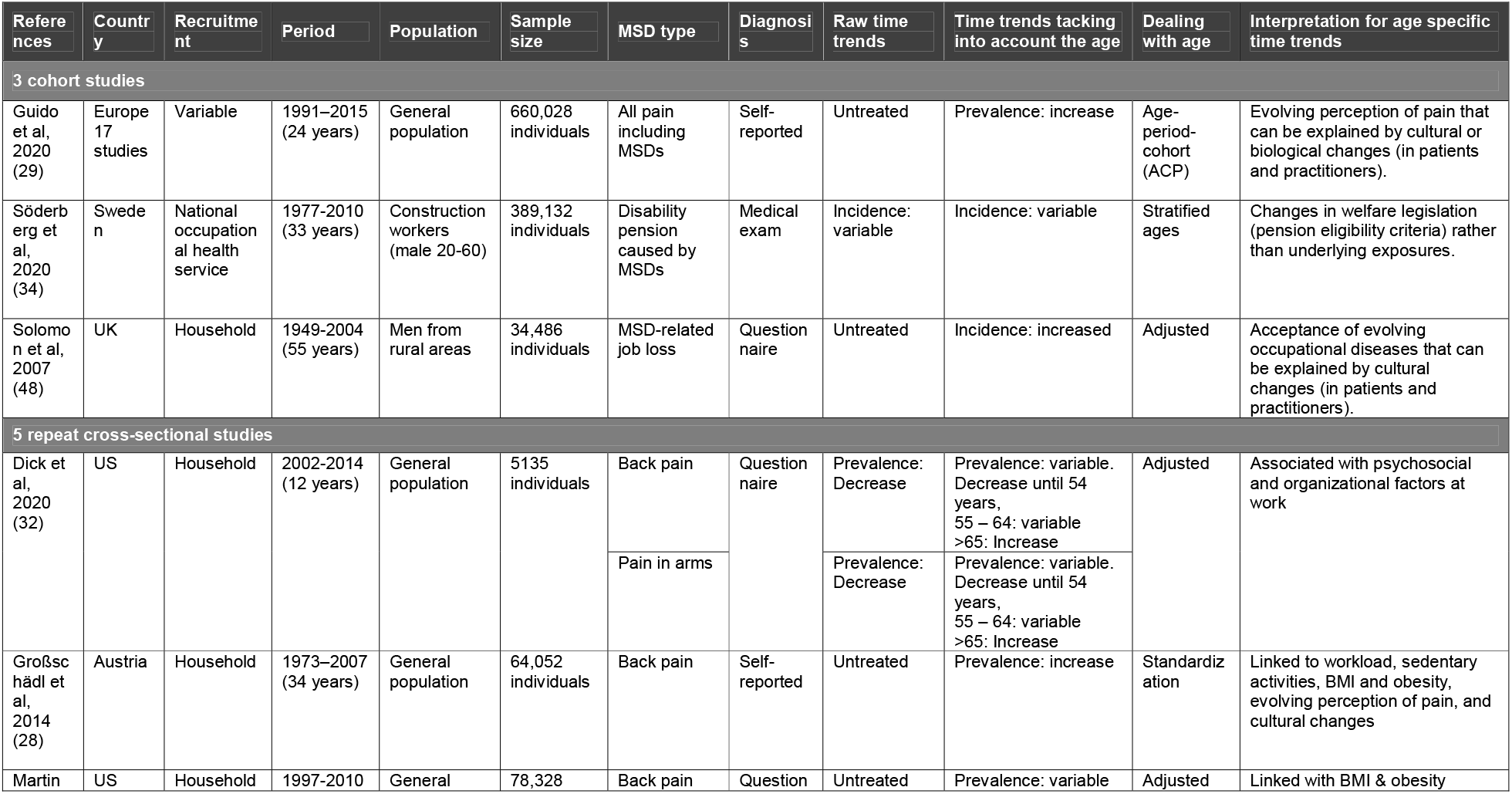
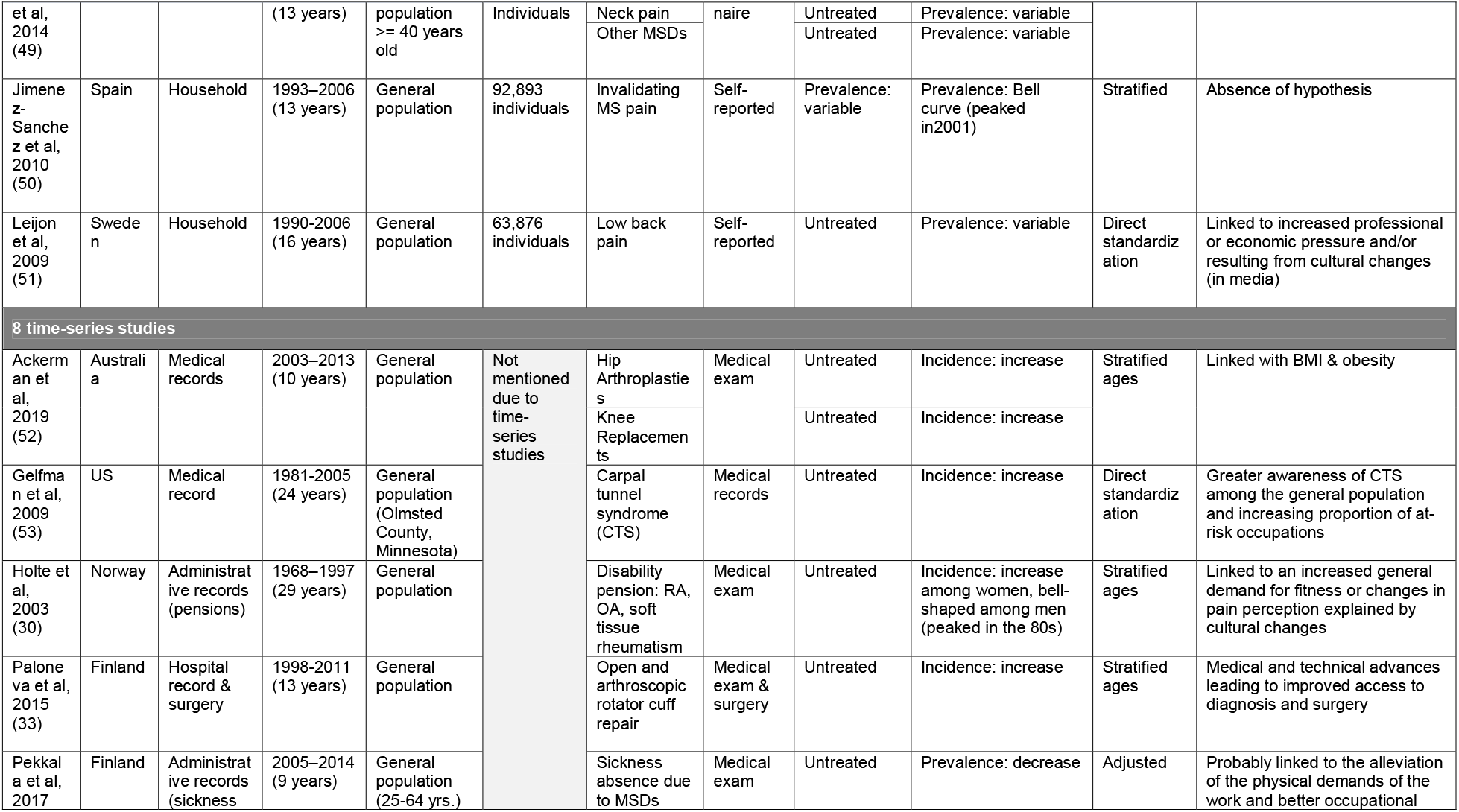
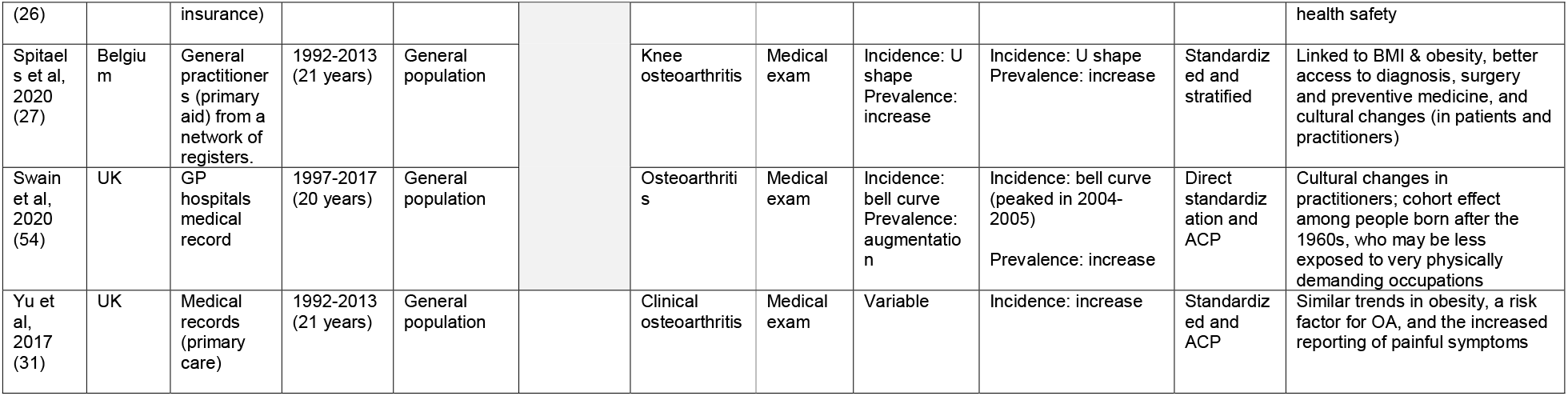
Summary of articles reporting temporal trends in MSD occurrence.

The recruitment of individuals for repeated cross-sectional studies was carried out from household-based sampling designs. Among cohort studies, 1 article relied on the recruitment of hospital-based participants, 1 recruited participants from occupational health records, and 1 recruited participants from previous surveys completed at home. Most of the time-series studies relied on hospital-based surveillance systems (5 out of 8 articles).

Among the articles included, 5 defined MSDs based on pain, and 11 defined them according to a disability. These 11 articles relying on a disability-related MSD definition were conducted in the Scandinavian countries, the United Kingdom, and Australia. Among those, the site of MSDs was not specified for 8 articles, 2 articles considered MSD affecting the inferior limbs and the back, and 1 considered MSD affecting the superior limbs.

### Risk of bias and quality of the studies

The studies selected were mainly carried out on the general working population, and the risk of selection bias was considered low or probably low for all studies. Overall, study participants were carefully selected based on a well-defined sampling strategy based on random selection from a national longitudinal or cross-sectional survey.

We considered MSDs based on medical diagnosis to be reliable. We classified both MSDs based on the medical diagnosis and/or disability at low risk of bias. MSDs defined based on pain were classified as a probably low risk of bias. For the studies which administered a questionnaire, we considered that they probably had low bias since it is a good method for detecting chronic pain and disability in the individuals recruited. Studies dealing with temporal trends in MSDs by controlling for age and then for other factors were considered at low risk of bias for the confounding factors. Studies not taking other potential confounders were considered likely to be at low risk of bias since here we are only looking at temporal trends in MSDs. In the included studies, most of the study authors did not declare a conflict of interest, nor did they receive any support from a company suggesting that there could be a financial interest in the results. Therefore, we assessed these studies as having a low risk of bias in this area. For the studies not clearly mentioning it in the paper, we verified that all the authors were affiliated with public (research) agencies or scientific institutions and, when this was the case, we considered that the studies had a low probability of bias. We did not identify any other biases and therefore assessed all studies as having a probable low risk of other biases.

Among 16 articles, 9 studies included either tests for temporal trends or confidence intervals for each value of MSD incidence or prevalence over time. These studies were considered to be of satisfactory quality (trends tests, or Chi-squared), or of probable satisfactory quality (95% CI for MSDs incidence and/or prevalence). The studies without statistical tests were considered to be of a probable unsatisfactory quality. In general, we did not identify studies where there was a high risk of bias, or where the quality was too low to justify an exclusion from the review (**Table 2**). Additional results regarding classification of bias and evaluation of statistical methods are provided in Supplementary material S4.

**Table 2:** Summary of risk of bias and quality across studies on temporal trends of MSDs

|  | Acker<br>man et<br>al,201<br>9<br>(52) | Dick et<br>al,<br>2020<br>(32) | Gelfman<br>et al,<br>2009<br>(53) | Großsch<br>ädl et al,<br>2014<br>(28) | Guido<br>et al,<br>2020<br>(29) | Holte<br>et al,<br>2003<br>(30) | Jimenez<br>-<br>Sanche<br>z et al,<br>2010<br>(50) | Leijon<br>et al,<br>2009<br>(51) | Martin<br>et al,<br>2014<br>(49) | Palonev<br>a et al,<br>2015<br>(33) | Pekkala<br>et al,<br>2017<br>(26) | Söderbe<br>rg et al,<br>2020<br>(34) | Solomo<br>n et al,<br>2007<br>(48) | Spitael<br>s et<br>al,<br>2020<br>(27) | Swain<br>et al,<br>2020<br>(54) | Yu et<br>al,<br>2017<br>(31) |
| --- | --- | --- | --- | --- | --- | --- | --- | --- | --- | --- | --- | --- | --- | --- | --- | --- |
| **Bias** |  |  |  |  |  |  |  |  |  |  |  |  |  |  |  |  |
| a. Bias in the<br>selection of<br>study<br>participants | L | L | L | L | L | L | L | L | L | L | L | L | PL | L | L | L |
| b. Bias due to<br>misclassificatio<br>n of MSDs | L | PL | L | PL | PL | L | PL | PL | PL | L | L | PL | PL | L | L | L |
| c. Bias due to<br>poor<br>consideration<br>of confounding<br>factors | L | L | L | L | L | L | L | L | L | L | L | PL | PL | L | PL | L |
| d. Bias due to<br>conflict of<br>interest | L | L | PL | L | L | PL | PL | L | PL | L | L | L | L | L | L | L |
| e. Other<br>biases | PL | PL | PL | PL | PL | PL | PL | PL | PL | PL | PL | PL | PL | PL | PL | PL |
| **Quality of the statistical tests** |  |  |  |  |  |  |  |  |  |  |  |  |  |  |  |  |
| Quality of the<br>statistical trend<br>tests | PUS | PSQ | PSQ | PUS | SQ | PUS | SQ | PSQ | SQ | PUS | SQ | PUS | PUS | SQ | PUS | PSQ |

Legend for the bias: low (L), probably low (PL), probably high (PH), high (H).
Legend for the quality of the statistical tests: satisfactory quality (SQ), probable satisfactory quality (PSQ), and probable unsatisfactory quality (PUS).

### Temporal trends of the incidence and prevalence of MSDs

Some studies simultaneously reported results for several MSDs and/or indicators (prevalence, incidence), such that the 16 included studies included a total of 23 results extracted for this review. Among these results, 9 used a definition of MSD based on pain and 14 based on impacts on work or social life. Among all results, 12 presented temporal trends in prevalence and 11 in incidence. The variability of the definitions of MSDs and body sites studied in the articles of our sample precludes meta-analysis and calculation of pooled estimates of temporal trends. Five studies controlled for age in time trends in MSDs by stratification, 4 articles by adjustment, 3 articles by standardizations, 3 articles by direct standardization, and 3 by age-period-cohort (see **Table 1**).

Among the 3 articles defining MSD prevalence based on disability (26), 1 showed that absences due to MSDs decreased over time after adjusting for age. Two articles reported an increase in MSDs over time, 1 of which reported increases in knee osteoarthritis (27), while the other reported increases in osteoarthritis (28) (**Figure 2.A**). Among the 9 results based on pain-related MSD definition, 7 showed non-monotonic change over time, and 2 reported increasing trends (Großschädl et al (28) for lower back pain, and Guido et al (29) for pain in all locations) (**Figure 2.B**). These results demonstrate heterogeneity in MSD time trends, including both increases and non-monotonic changes (**Table 1**).

**Figure 2:**
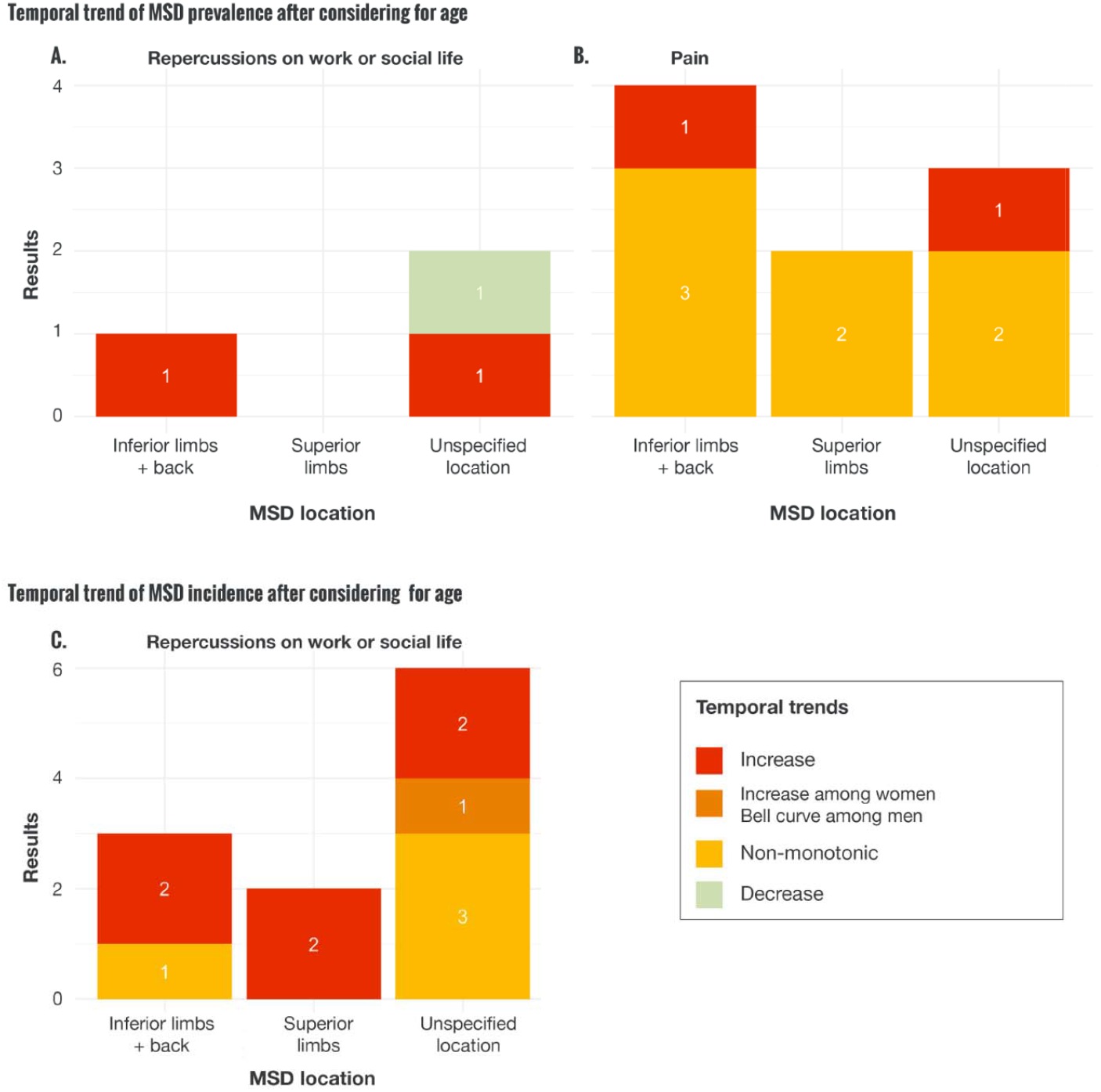
Temporal trends of the incidence and prevalence of MSDs according to their location and severity. **A**. Temporal trend of the prevalence of MSDs inducing repercussions on work or social life with age considerations. **B**. Temporal trend of the prevalence of pain with age considerations. **C**. Temporal trend of the incidence of MSDs inducing repercussions on work or social life with age considerations Unspecified location refers to MSDs that were not associated with a specified body site.

The temporal evolution of the incidence of MSDs causing disability also tended to increase or vary according to their site. Among the 6 articles not specifying MSD location, 3 showed variable trends, two reported increases over time and 1 (30) reported sex-specific results, with an increase in women and a bell curve for men. In addition, two articles reported an increase over time in MSDs located in superior limbs. For the inferior limbs and the back, two articles showed an increase in MSDs, and 1 article reported a variable evolution of MSDs (**Figure 2.C** and **Table 1**).

### Synthesized evidence

We note that few articles analyzed temporal variations in MSDs while controlling for age.

Controlling or not controlling for age may produce divergent pictures of temporal trends in specific MSDs. In the article by Yu et al (31), the raw data suggested non-monotonic changes in osteoarthritis incidence over time, whereas increased incidence was observed when standardizing for age. In the article by Dick et al (32), the unadjusted prevalence of back and hand pain decreased over time. After adjusting for age, decreased prevalence was still observed among people < 55 years, but variable time trends were observed for those aged 55-64 years, and an increase was observed for those ≥ 65 years. Thus, these results highlight potentially distinct impacts of age and time in the occurrence of MSDs, at least for older age categories.

Fifteen articles suggested that factors other than ageing could explain temporal MSD trends (**Table 1**). Regardless of the trends observed, most articles hypothesized a link between cultural changes around the perception of pain (in both caregivers and patients), and a better knowledge of pathologies with improvements in detection and treatment techniques (29-33). We also noticed that 5 articles related the trends they reported in MSDs to similar temporal trends in obesity and body mass index. Additionally, Dick et al (32) suggested that changes in psychosocial and organizational factors at work could explain the non-monotonic trends that they observed between 2002 and 2014. Söderberg et al (34) suggested that the non-monotonic trends they reported reflected changes in disability eligibility criteria rather than in the underlying exposures. Finally, the only article which reported a decrease in the instances of sick leave due to MSD (26) explained it by a probable reduction in the physical demands of work and better health and safety at work.

## DISCUSSION

This literature review identified a limited number of articles reporting temporal trends in MSDs while controlling for age. Study duration ranged from 10 to 55 years, which allowed for longitudinal analysis of MSD occurrence. Temporal trends in MSDs varied according to the site of the MSDs, the criteria used to define MSDs (either associated with pain and/or a disability), and the indicator used (prevalence or incidence). We observed temporal heterogeneity in the occurrence of MSDs considered, with mainly non-monotonic or increasing trends reported. Of note, based on studies reporting both crude and age-controlled indicators, we observed that accounting or not accounting for age could lead to diverging temporal trends, at least among the highest age categories.

This literature review identified some important gaps and residual uncertainty in the evidence currently available. First, although our inclusion criteria were broad, the systematic review only identified studies conducted in Western, high-income countries: USA, Europe (especially Scandinavian countries), and Australia. This lack of evidence considering the burden of MSDs and their socio-economic implications does not allow us to provide an interpretation of the evolution of MSDs among the global working population (35).

Occurrence of the different groups of MSDs considered in this review (pain vs. disability) varied over time depending on the indicator considered (prevalence vs. incidence). To diagnose the occurrence of MSDs, several scales allow for quickly and easily assessing pain intensity (visual analog pain scale, simple numeric scale, simple verbal scale) (36, 37). It is important to note, however, that although the validity of these diagnostic tests is comparable in educated patients, those who are less or uneducated may be led to answer differently. These scales do not allow for a complete assessment of the pain component, but they can allow for repeated self-assessments since they are very quick to complete. It is also possible that the pain reported by patients responds more quickly to changes in working conditions or other factors (such as cultural changes) than do longer disabling pathologies. Therefore, we must remain cautious about our interpretations of temporal changes in the occurrence of MSDs, depending on whether the observed outcome relates to self-reported pain or more disabling pathologies diagnosed by doctors (38). Pain classification measurements must therefore include aspects such as the severity, frequency, and intensity of pain as well as measurements of changes in working conditions (39).

We hypothesize that observed heterogeneity in temporal trends of MSD occurrence results from temporal heterogeneity in the evolution of MSD risk factors in different populations. In most of the countries covered by this review, a fundamental change in the tertiarization of work has been observed, resulting in an overall reduction in occupational physical constraints (40, 41). However, a reduction in MSD is not systematically expected from decreased exposure to biomechanical factors. The analyses from the ESTEV survey show in particular that the viscoelastic nature of periarticular soft tissues can also play a role in the occurrence of low back pain (42). Thus, prolonged exposure from carrying heavy loads can potentially cause an irreversible deformation of these tissues (“memory of the exposure” or “creep phenomenon”), which may explain the fact that, in older age groups, some MSDs have not decreased despite decreased biomechanical exposures. Moreover, a decrease in occupational physical constraints may have arisen concomitantly with an increase in work-related mental load, which can also play a significant role in the occurrence of MSDs (8, 43).

The main limitation of this review results from the fact that we exclusively searched electronic bibliometric databases of scientific literature. This means that we did not consult the gray literature or governmental reports on MSDs that were not peer-reviewed by external readers. Another limitation is that, since we used the generic term musculoskeletal disorders/disease as a keyword, it is possible that we missed articles on specific MSDs that did not mention the term MSD in the abstract or key terms. Lastly, variability in MSD definitions and body sites among our study sample prevented us from conducting a meta-analysis and computing pooled estimates of time trends.

Finally, we do not have studies capturing MSD data during the health crisis linked to the COVID-19 pandemic. This sanitary situation could possibly be at the origin of the evolution and emergence of certain professions which can potentially be at the origin of changes in the occurrence of MSDs (telework and bad postures, sedentary habits, intensification of work, work on task linked to a digital platform, increased deliveries carrying heavy loads at reduced times, stress, etc.) (44-46). In the future, longitudinal data that can capture this information could be an interesting addition to the interpretation and understanding of the occurrence of MSDs over time (47).

## CONCLUSION

To our knowledge, this is the first systematic search and review of studies reporting on MSD occurrence while accounting for the key confounding impact of age. Our findings suggest disparity in the literature regarding the temporal evolution of MSD occurrence, depending on the site of the MSD and whether accounting for MSDs defined by scales of self-reported pain or disability. Overall, studies controlling for age reported either non-monotonic changes or increases in MSD occurrence over time. Factors other than ageing that have also been suggested to underlie temporal changes in MSD occurrence include changes in obesity, occupational and cultural exposures, and pain tolerance. The current body of evidence, however, highlights residual uncertainties, especially given the limited number of articles on this subject and the fact that we only found articles in wealthy countries. Notably, this review demonstrates the type of research and data that are lacking to anticipate the temporal trends in the MSDs occurrence, which is an important question in terms of prevention. We also showed that in the included articles, the temporal trends of MSDs varied mainly between increase and non-monotonic changes depending on their site, severity, and age.

## Data Availability

In this literature review we use electronic bibliographic databases for studies published between 1990 and 2020: Medline, ScienceDirect, Wiley, and Web of Science

## ACKNOWLEDGMENTS

The authors are responsible for the opinions expressed in this article, and they do not necessarily represent the views, decisions, or policies of the institutions with which they are affiliated. We also would like to thank David R M Smith for his precious help and careful proofreading of our work.

