## supplementary materials for "Can we distinguish the roles of demographic and temporal changes in the incidence and prevalence of musculoskeletal disorders? A systematic search and review"

**S1: Supplementary material - PRISMA 2020 checklist**

| **Section and Topic** | **Item #** | **Checklist item** | **Location where item is reported** |
| --- | --- | --- | --- |
| **TITLE** | | | Line 1-3 |
| Title | 1 | Identify the report as a systematic review. |  |
| **ABSTRACT** | | |  |
| Abstract | 2 | See the PRISMA 2020 for Abstracts checklist. | Lines 6-25 |
| **INTRODUCTION** | | |  |
| Rationale | 3 | Describe the rationale for the review in the context of existing knowledge. | Line 34-59 |
| Objectives | 4 | Provide an explicit statement of the objective(s) or question(s) the review addresses. | Line 61-65 |
| **METHODS** | | |  |
| Eligibility criteria | 5 | Specify the inclusion and exclusion criteria for the review and how studies were grouped for the syntheses. | Lines 81-101 |
| Information sources | 6 | Specify all databases, registers, websites, organisations, reference lists and other sources searched or consulted to identify studies. Specify the date when each source was last searched or consulted. | Lines 75-80 |
| Search strategy | 7 | Present the full search strategies for all databases, registers and websites, including any filters and limits used. | Lines 75-101 |
| Selection process | 8 | Specify the methods used to decide whether a study met the inclusion criteria of the review, including how many reviewers screened each record and each report retrieved, whether they worked independently, and if applicable, details of automation tools used in the process. | Line 103-107 |
| Data collection process | 9 | Specify the methods used to collect data from reports, including how many reviewers collected data from each report, whether they worked independently, any processes for obtaining or confirming data from study investigators, and if applicable, details of automation tools used in the process. | Line 109-113 |
| Data items | 10a | List and define all outcomes for which data were sought. Specify whether all results that were compatible with each outcome domain in each study were sought (e.g. for all measures, time points, analyses), and if not, the methods used to decide which results to collect. |  |
|  | 10b | List and define all other variables for which data were sought (e.g. participant and intervention characteristics, funding sources). Describe any assumptions made about any missing or unclear information. |  |
| Study risk of bias assessment | 11 | Specify the methods used to assess risk of bias in the included studies, including details of the tool(s) used, how many reviewers assessed each study and whether they worked independently, and if applicable, details of automation tools used in the process. | Line 115-124 |
| Effect measures | 12 | Specify for each outcome the effect measure(s) (e.g. risk ratio, mean difference) used in the synthesis or presentation of results. | Line 126-138 |
| Synthesis methods | 13a | Describe the processes used to decide which studies were eligible for each synthesis (e.g. tabulating the study intervention characteristics and comparing against the planned groups for each synthesis (item #5)). | Not applicable here |
|  | 13b | Describe any methods required to prepare the data for presentation or synthesis, such as handling of missing summary statistics, or data conversions. |  |
|  | 13c | Describe any methods used to tabulate or visually display results of individual studies and syntheses. | Line 140-144 & 223-224 |
|  | 13d | Describe any methods used to synthesize results and provide a rationale for the choice(s). If meta-analysis was performed, describe the model(s), method(s) to identify the presence and extent of statistical heterogeneity, and software package(s) used. |  |
|  | 13e | Describe any methods used to explore possible causes of heterogeneity among study results (e.g. subgroup analysis, meta-regression). | Not applicable here |
|  | 13f | Describe any sensitivity analyses conducted to assess robustness of the synthesized results. |  |
| Reporting bias assessment | 14 | Describe any methods used to assess risk of bias due to missing results in a synthesis (arising from reporting biases). | Not applicable here |
| Certainty assessment | 15 | Describe any methods used to assess certainty (or confidence) in the body of evidence for an outcome. |  |
| **RESULTS** | | |  |
| Study selection | 16a | Describe the results of the search and selection process, from the number of records identified in the search to the number of studies included in the review, ideally using a flow diagram. | Line 147-154 + figure 1 |
|  | 16b | Cite studies that might appear to meet the inclusion criteria, but which were excluded, and explain why they were excluded. | Not applicable here |
| Study characteristics | 17 | Cite each included study and present its characteristics. | Line 156-176 |
| Risk of bias in studies | 18 | Present assessments of risk of bias for each included study. | Line 178-206  + Table 2 |
| Results of individual studies | 19 | For all outcomes, present, for each study: (a) summary statistics for each group (where appropriate) and (b) an effect estimate and its precision (e.g. confidence/credible interval), ideally using structured tables or plots. | Table1 |
| Results of syntheses | 20a | For each synthesis, briefly summarise the characteristics and risk of bias among contributing studies. | Line 184-197 |
|  | 20b | Present results of all time trends syntheses conducted. | Line 208-231 |
|  | 20c | Present results of all investigations of possible causes of heterogeneity among study results. | Line 223-224 |
|  | 20d | Present results of all sensitivity analyses conducted to assess the robustness of the synthesized results. | Not applicable here |
| Reporting biases | 21 | Present assessments of risk of bias due to missing results (arising from reporting biases) for each synthesis assessed. | Not applicable here |
| Certainty of evidence | 22 | Present assessments of certainty (or confidence) in the body of evidence for each outcome assessed. | Line 233-254 |
| **DISCUSSION** | | |  |
| Discussion | 23a | Provide a general interpretation of the results in the context of other evidence. | Line 257-265 |
|  | 23b | Discuss any limitations of the evidence included in the review. | Line 267-300 |
|  | 23c | Discuss any limitations of the review processes used. | Line 302-308 |
|  | 23d | Discuss implications of the results for practice, policy, and future research. | Line 310-316 |
| **OTHER INFORMATION** | | |  |
| Registration and protocol | 24a | Provide registration information for the review, including register name and registration number, or state that the review was not registered. | Line 69-73 |
|  | 24b | Indicate where the review protocol can be accessed, or state that a protocol was not prepared. |  |
|  | 24c | Describe and explain any amendments to information provided at registration or in the protocol. |  |
| Support | 25 | Describe sources of financial or non-financial support for the review, and the role of the funders or sponsors in the review. | Line 417-420 |
| Competing interests | 26 | Declare any competing interests of review authors. | Line 422-424 |
| Availability of data, code and other materials | 27 | Report which of the following are publicly available and where they can be found: template data collection forms; data extracted from included studies; data used for all analyses; analytic code; any other materials used in the review. | Supplementary material S2 |

*From:* Page MJ, McKenzie JE, Bossuyt PM, Boutron I, Hoffmann TC, Mulrow CD, et al. The PRISMA 2020 statement: an updated guideline for reporting systematic reviews. BMJ 2021;372:n71. doi: 10.1136/bmj.n71

For more information, visit: <http://www.prisma-statement.org/>

**Supplementary material 2:** Search strategy used for each database, with the detail of the algorithms of keywords used, the number of results per database and the total number of articles identified

| Database | Key words algorithms | Number of articles | Total per database | Total articles |
| --- | --- | --- | --- | --- |
| Medline | 1. "Musculoskeletal diseases*"[All Fields] AND ("temporal trends"[All Fields] OR "time trends"[All Fields] OR "over time"[All Fields] ) AND ("incidence"[All Fields] OR "prevalence"[All Fields]) AND ("1990/01/01"[PubDate] : "2020/01/30"[PubDate]) | 1,243 | 1,552 | 2,680 |
|  | 2. "Musculoskeletal disorders*"[All Fields] AND ("temporal trends"[All Fields] OR "time trends"[All Fields] ) AND ("incidence"[All Fields] OR "prevalence"[All Fields]) AND ("1990/01/01"[PubDate] : "2020/01/30"[PubDate]) | 309 |  |  |
| Science Direct | 1. ("Musculoskeletal disease" OR "Musculoskeletal diseases") AND ("temporal trends" OR "time trends"OR "over time") AND ("incidence“ OR "prevalence") and select by hand 1990-2020 | 547 | 618 |  |
|  | 2. ("Musculoskeletal disorders"OR"Musculoskeletal disorders") AND ("temporal trends" OR "time trends"OR "over time") AND ("incidence“ OR "prevalence") and select by hand 1990-2020 | 71 |  |  |
| Wiley | 1. ("Musculoskeletal disease"OR"Musculoskeletal diseases") AND ("temporal trends" OR "time trends"OR "over time") AND ("incidence“ OR "prevalence") and select by hand 1990-2020 | 379 | 472 |  |
|  | 2. ("Musculoskeletal disorders"OR"Musculoskeletal disorders") AND ("temporal trends" OR "time trends"OR "over time") AND ("incidence“ OR "prevalence") and select by hand 1990-2020 | 93 |  |  |
| Web of Science | 1. ("Musculoskeletal disease"OR"Musculoskeletal diseases") AND ("temporal trends" OR "time trends"OR "over time") AND ("incidence“ OR "prevalence") and select by hand 1990-2020 | 18 | 38 |  |
|  | 2. ("Musculoskeletal disorders"OR"Musculoskeletal disorders") AND ("temporal trends" OR "time trends"OR "over time") AND ("incidence“ OR "prevalence") and select by hand 1990-2020 | 20 |  |  |

**Supplementary material S3:** MSD definition by article

| **Reference** | **Country** | **Definition of the MSD reported in each article** |
| --- | --- | --- |
| Ackerman et al,2019  (1) | Austria | Primary total knee (TKR) and hip (THR) arthroplasties performed for osteoarthritis. See the link below to view the source they used for this definition: <https://aoanjrr.sahmri.com/documents/10180/689619/Hip%2C+Knee+%26+Shoulder+Arthroplasty+New/6a07a3b8-8767-06cf-9069-d165dc9baca7> |
| Dick et al, 2020 (2) | US | In this study ,MSD definitions were collected by questionnaires supervised by an interview in a survey. The outcomes were the prevalence of back pain and pain in arms through a yes/no response to ‘‘In the past 12 months, have you had back pain every day for a week or more?’’ and ‘‘In the past 12 months, have you had pain in the hands, wrists, arms, or shoulders every day for a week or more?’’ |
| Gelfman et al, 2009  (3) | US | CTS: entrapment of the median nerve in the carpal tunnel, which is formed by the flexor retinaculum and carpal bones; this syndrome may be associated with repetitive occupational trauma, wrist injuries, amyloid neuropathies, rheumatoid arthritis, acromegaly, pregnancy, and other conditions; symptoms include burning pain and paresthesias affecting the ventral surface of the hand and fingers which may radiate proximally; altered sensation in the distribution of the median nerve and thenar atrophy may occur http://www.icd9data.com/2015/Volume1/320-389/350-359/354/354.0.htm |
| Großschädl et al, 2014  (4) | Austria | In each survey, the presence of back pain was questioned. When collecting data from the Microcensus surveys (1973, 1983, 1991, 1999), participants were asked if they suffered from back pain at the time of the survey. If they have suffered from the disorder in the past 12 months. Surveys have used different definitions to identify back pain. In the last survey, the 12-month prevalence of back pain was collected, while in the first four Microcensus surveys, the point prevalence was measured. Despite the different collection methods of Microcensus and AT-HIS, it has been reported that back pain is often chronic and that there is not much difference in the prevalence of back pain at the time of investigation or rather within 12 months. The back pain data from the AT-HIS 2006–07 did not appear obvious and were similar and roughly matched the data from the Microcensus surveys. Therefore, the effect of the different definitions is probably minimal and reflects only a slight change in the prevalence of back pain over the study period. |
| Guido et al, 2020 (5) | Europe - 17 different countries | Depending on the country, the definition varies: Pain is included in the state of health and functional limitations. It was measured in 14 of the 17 studies, with different approaches. Some studies, for example, *Collaborative Research on Aging in Europe and Longitudinal Study on Health and Retirement in China* have approached it in terms of pain severity (e.g. none, mild, moderate, severe, extreme or other similar formats). Other studies, e.g. the Australian Longitudinal Study on Aging and the European Health, Aging and Retirement Survey dichotomously addressed the presence of pain (e.g. yes or no), sometimes addressing the idea that pain is 'often felt', as in the Irish Longitudinal Study on Aging. The harmonization procedure aims to generate inferentially equivalent content across studies to make the content of the variables collected in different studies uniform. For the pain variable, the content was "self-reported pain experienced at the time of the interview", and the modality of the variable was dichotomous. |
| Holte et al,  2003 (6) | Norway | Rheumatoid arthritis (RA): is a form of arthritis that causes pain, swelling, stiffness, and loss of function in the joints. It can affect any joint, but it is common in the wrist and fingers. More women than men suffer from rheumatoid arthritis. It often begins between the ages of 25 and 55. You may only have the disease for a short time, or the symptoms may come and go. The severe form can last a lifetime. Rheumatoid arthritis is different from osteoarthritis, the common arthritis that often occurs with age. RA can affect parts of the body in addition to the joints, such as your eyes, mouth, and lungs. RA is an autoimmune disease, which means arthritis results from your immune system attacking your body's own tissues. No one knows what causes rheumatoid arthritis. Genetics, the environment, and hormones could all contribute. Treatments include medicine, lifestyle changes, and surgery. These can slow or stop joint damage and reduce pain and swelling. NIH: National Institute of Arthritis, Musculoskeletal and Skin Diseases |
| Jimenez-Sanchez et al, 2010 (7) | Spain | Musculoskeletal disorders: subjects who have suffered from self-reported musculoskeletal pain (bone, spinal, or joint pain). If during the previous 2 weeks pain induces a decrease or limits the main work activity or free time activity by at least half a day, then symptoms are classified as disabling musculoskeletal pain. |
| Leijon et al, 2009 (8) | Sweden | The questionnaires included a question on low back pain, modified from the Standardized Nordic Questionnaire, with a recall period of 12 months (1990 and 1994) or 6 months (1998, 2002 and 2006): the last six (twelve) months? '' The questions had identical alternative answers in each of the five surveys: "No, never", "Yes, a few days in the last six (twelve) months", "Yes, a few days a month", "Yes, a few days a week” and “Yes, every day.” From a clinical point of view and for the purposes of this study, low back pain was defined as pain a few days a week or every day. |
| Martin et al, 2014 (9) | Finland | Questionnaire on 9 physical functions, pain that induces difficulty in: bending, bending, or kneeling, standing for 2 hours, pushing or pulling a large object; walk a quarter of a mile; climb ten steps; seated 2 hours; lift and carry ten pounds; reach above the head; and grab small items. |
| Paloneva et al, 2015  (10) | Finland | Arthroscopic open repair of the rotator cuff according to the Nomesco Classification of Surgical Procedures (Finnish version). |
| Pekkala et al, 2017  (11) | Finland | The diagnostic causes have been classified according to the main chapters of ICD-10: musculoskeletal diseases (M00 - M99). See the list of diseases on the WHO website below. Chapter definition: Diseases of the osteoarticular system, the muscles and connective tissue <https://icd.who.int/browse10/2008/fr#/XIII> |
| Söderberg et al, 2018  (12) | Sweden | Musculoskeletal diagnosis according to the definition of ICD10 code M00-M99. Chapter definition: Diseases of the osteoarticular system, muscles and connective tissue. See the link below for to go further in the definition with the source they used <https://icd.who.int/browse10/2008/fr#/XIII> |
| Solomon et al, 2007  (13) | United Kingdom | Questionnaire distributed, with the question concerning MSDs: "Have you ever left or abandoned a job (including jobs held for less than a year) because of a health problem?" Then question on muscle pain and recoding in MSD. |
| Spitaels et al, 2020 (14) | Belgium | Osteoarthritis of the knee: Criteria: Either imaging with a characteristic appearance; either a joint disorder that has progressed for at least three months, without constitutional symptoms comprising three or more of the following three signs: intermittent swelling, crepitus, stiffness or limitation of movement, speed of sedimentation, rheumatoid tests, normal uric acid; over 40 years. Includes: knee osteoarthritis secondary to dysplasia; knee osteoarthritis secondary to trauma. See the link below to view the source they used for this definition: <https://www.hetop.eu/hetop/3CGP/en/?rr=CIP_D_L91&q=CIP_D_L91#rr=CIP_D_L90&q=CIP_D_L90> |
| Swain et al,  2020  (15) | United Kingdom | The osteoarthritis incident was defined as the first diagnosis of osteoarthritis in each year of study. Prevalent osteoarthritis was defined as having a diagnosis of osteoarthritis on July 1 of each year of study. Read codes were used: a medical coding system for clinical terms used by the National Health Services (NHS), United Kingdom. The Read code list available (www.keele.ac.uk/mrr) to identify people with osteoarthritis diagnosed by general practitioners (GPs) has been adapted according to the inclusion and exclusion criteria of the study. Two types of osteoarthritis were excluded (acromioclavicular and sternoclavicular joints), due to the possible low precision of the diagnosis at the level of these joints and the expected incidence is very low. The codes obtained from the given website were previously mapped to the ICD-10 codes. |
| Yu et al,  2017 (16) | United Kingdom | Two definitions of osteoarthritis: 1) Defined cases having had at least 1 consultation with a recorded diagnosis of osteoarthritis or, at least 1 consultation with a recorded peripheral joint pain symptom affecting the knee, hip and hand/wrist likely to reflect osteoarthritis (clinical osteoarthritis); 2) Cases defined more narrowly as having at least 1 consultation with a recorded diagnosis of osteoarthritis. |

**Supplementary material 4:** Detail of bias risk and quality in statistical analysis

An important step in systematic literature review methods is to assess the risk of bias of individual studies. Therefore, we adapted the methodology of two tools usually used : the RoB -SPEO tool (17) and the Navigation Guide's evidence quality assessment tool inspired by the article by Alexis Descatha et al, 2018 (appendix H) (18).

From the RoB-SPEO criteria (17) we used bias: 1) in selection of participants into the study, 2) due to misclassification of MSD, 3) due to conflict of interest, and 4) other bias. Thus, we considered 4 biases among 8 since the other bias was not relevant because of their specificity to exposures and measures of risk factors and in our study we only assessed the time trends in MSDs (reminder of the biases that have not been retained because they are not relevant here: bias due to a lack of blinding of study personnel, bias due to incomplete exposure data, bias due to selective reporting of exposures and bias due to differences in numerator and denominator).

In the Navigator guide described in the Alexis Descatha et al, 2018 (appendix H) article (18), most of the criteria were close to RoB-SPEO but we added the relative criterion on confounding factors which was consistent with the needs of our study (bias due to poor consideration of confounding factors).

Classifications of each bias for the selected articles are provided below.

**S4.a:** Selection bias: potential bias resulting from study groups not adequately representing the population of interest.

- Low: the target population represented the whole working population or general population.
- Probably low: there was insufficient information about participant selection to permit a judgment of low risk of bias, but the worker population used in the study was specific to one category of worker (eg: farmer, worker, executives, etc.).
- Probably high there was insufficient information about participant selection to permit a judgment of high risk of bias, but there is indirect evidence which suggests that inclusion/exclusion criteria, recruitment and enrollment procedures, and participation and follow-up rates were inconsistent across groups, as described by the criteria for a judgment of high risk of bias.
- High: participant selection was based on voluntary participation, or there were indications from descriptions of the source population that risk of selection effects were substantial, whether due to issues in inclusion/exclusion criteria, recruitment and enrolment procedures, participation and follow-up rates, or data on the distribution of relevant study sample and population characteristics.

| **Bias** | **References** | **Ladder** | **Information found in each article related to the bias** |
| --- | --- | --- | --- |
| Bias in selection of participants into the study | Ackerman et al., 2019 | Low | Anonymized individual data obtained from the Australian Orthopaedic Association National Joint Replacement Registry (AOANJRR). Data collected from all public and private hospitals performing joint replacements. |
|  | Dick et al., 2020 | Low | Data collected as part of the biannual General Social Survey (GSS) conducted by the National Opinion Research Center (NORC) at the University of Chicago. This survey is described as nationally representative and was conducted to track societal changes and study American society. The Quality of Life at Work (QWL) module is the module that has been added to collect nationally representative opinions on working life. |
|  | Gelfman et al., 2009 | Low | Data from a medical registry (from the Rochester Epidemiology Project) in the population of Olmsted County (US). |
|  | Großschädl et al., 2014 | Low | Data from cross-sectional health surveys conducted by random selection. |
|  | Guido et al., 2020 | Low | Data from existing international longitudinal cohorts (ATHLOS project) related to health and aging. The harmonized data set includes participant records from 17 different studies. |
|  | Holte et al., 2003 | Low | Data file from the National Insurance Administration (NIA) -> disability pension. |
|  | Jimenez-Sanchez et al., 2010 | Low | Data obtained from the Spanish national health system via a permanent personal home interview that examines a nationally representative sample of the non-institutionalized civilian population residing in major family dwellings in Spain. |
|  | Leijon et al., 2009 | Low | Cross-sectional survey data with random drawing and stratification according to the population structure of Stockholm. The diagnoses included in the analyses of hospitalization were from the International Classification of Diseases versions 9 and 10 (ICD 9 and ICD 10). |
|  | Martin et al., 2014 | Low | National data from the National Health Interview Survey (NHIS), which is an annual nationally representative survey of the non-institutionalized population of the United States. |
|  | Paloneva et al., 2015 | Low | National data from the Finnish Hospital Discharge Register (FHDR) for the period of 1998-2011. |
|  | Pekkala et al., 2017 | Low | Data based on a nationally representative 70% random sample of Finnish residents. The sample data was unbalanced across years since individuals could be included in the sample each year (or enter and exit the data set). |
|  | Söderberg et al., 2018 | Low | Data from the national occupational health service listing the basic clinical examinations of workers every five years (cohort). Possible identification of subjects who have obtained a disability pension. |
|  | Solomon et al., 2007 | Probably low | Study population including men working in agriculture at the time of the national census. |
|  | Spitaels et al., 2020 | Low | Intego database including data extracted from electronic health records of general practitioners, all using Medidoc medical software. |
|  | Swain et al., 2020 | Low | Data from general practice electronic medical records which can be generalized to the general UK population. |
|  | Yu et al., 2017 | Low | Longitudinal data from the UK Clinical Practice Research Datalink (CPRD) which contains primary care records from GP surgeries covering around 7% of the UK population. |

**S4.b:** Bias of misclassification: the potential biases linked to a misclassification of MSDs .

- Low: the diagnosis was made by a medical exam
- Probably low: the diagnosis was made by and/or via an interview or auto-questionnaire.
- Probably high: information on the diagnosis was insufficient to judge a high risk of bias, but there was indirect evidence suggest that the symptom identification method criteria, were inconsistent between groups, the bias was considered to be probably high. For example, when the questionnaires distributed according to the groups were not the same each year.
- High: If it was proven that the diagnosis was not an element of the study design capable of introducing a risk of bias into the study, the bias was considered to be high.

| **Bias** | **References** | **ladder** | **Information found in each article related to the bias** |
| --- | --- | --- | --- |
| Bias due to misclassification of MSDs | Ackerman et al., 2019 | Low | Medical diagnostics |
|  | Dick et al., 2020 | Probably low | The survey is a 90-minute, face-to-face survey administered to randomly selected non-institutionalized American adults aged 18 and over. Primary outcomes were prevalence of back pain and arm pain with a yes/no response to "In the past 12 months, have you had back pain every day for a week or more?" and ''During the past 12 months, have you had pain in your hands, wrists, arms or shoulders every day for a week or more?” |
|  | Gelfman et al., 2009 | Low | Medical diagnostics |
|  | Großschädl et al., 2014 | Probably low | Interview via a questionnaire on the health of the individuals surveyed |
|  | Guido et al., 2020 | Probably low | Pain has been approached differently in different countries: sometimes dichotomously (presence of pain yes or no); and sometimes the idea that pain is "often felt" has been gradually defined. There was a harmonization procedure that aimed to generate inferentially equivalent content between the studies to make the content of the variables collected in the different studies uniform. |
|  | Holte et al., 2003 | Low | Medical diagnostics |
|  | Jimenez-Sanchez et al., 2010 | Probably low | Questionnaires on pain |
|  | Leijon et al., 2009 | Probably low | Questionnaires on pain |
|  | Martin et al., 2014 | Probably low | Questionnaires on pain |
|  | Paloneva et al., 2015 | Low | Medical diagnostics |
|  | Pekkala et al., 2017 | Low | Medical diagnostics |
|  | Söderberg et al., 2018 | Probably low | Disability pension following a medical diagnosis |
|  | Solomon et al., 2007 | Probably low | Questionnaire on job losses due to MSD or MSD proxies |
|  | Spitaels et al., 2020 | Low | Medical diagnostics |
|  | Swain et al., 2020 | Low | Medical diagnostics |
|  | Yu et al., 2017 | Low | Medical diagnostics |

**S4.c:** Bias due to incorrectly taking confounding factors into account:

Regarding bias due to a poor consideration of confounding factors, this criterion had to be adapted to our study. In our study, we sought to review the articles which deal with the temporal evolution of MSDs by taking age into account to see whether it is possible to distinguish the effect of age from that of time in the occurrence of these pathologies. Therefore, since in our selection procedure, we only selected articles dealing with temporal trends in MSDs by age (and not only raw trends) we do not risk having selected articles for which there will be high or probably high risk of bias (absence of the age factor). We therefore defined 2 categories of evaluations:

Low: when, in addition to the age factor, other confounding factors, the bias was considered to be low. These factors provided a more detailed analysis of temporal trends

Probably low: when only the age factor has been considered, the bias was defined as probably low. The age factor allows to take into account the effect of demographics in the temporal trends of the occurrence of MSDs.

| **Bias** | **References** | **ladder** | **Information found in each article related to the bias** |
| --- | --- | --- | --- |
| Bias due to poor consideration of confounding factors | Ackerman et al., 2019 | Low | National population projections, stratified by age and sex, were obtained from the Australian Bureau of Statistics (ABS). These projections were based on national census data and a series of assumptions about future fertility, life expectancy and migration. |
|  | Dick et al., 2020 | Low | Controlling the age, sex, mental and physical health, sleeping problems, physical factors, hand movements, psychosocial and organizational risks, etc. |
|  | Gelfman et al., 2009 | Low | Controlling the age and sex. |
|  | Großschädl et al., 2014 | Low | Each survey sample was weighed according to sex, age and region to ensure representation of the Austrian population. Data analysis for this study was limited to adults. |
|  | Guido et al., 2020 | Low | Controlling the age, sex, and time. |
|  | Holte et al., 2003 | Low | Controlling the age, sex, and time. |
|  | Jimenez-Sanchez et al., 2010 | Low | Taking socio-demographic variables into account. |
|  | Leijon et al., 2009 | Low | Controlling the age and sex. |
|  | Martin et al., 2014 | Low | Controlling the age, sex, and proxy response to the NHIS family basic questionnaire. |
|  | Paloneva et al., 2015 | Low | Controlling the age, sex. |
|  | Pekkala et al., 2017 | Low | Controlling the age, sex. |
|  | Söderberg et al., 2018 | Probably low | Consideration of age but no other potential confounding factors. |
|  | Solomon et al., 2007 | Probably low | Consideration of age but no other potential confounding factors. |
|  | Spitaels et al., 2020 | Low | Controlling the age, sex. |
|  | Swain et al., 2020 | Probably low | Consideration of age but no other potential confounding factors. |
|  | Yu et al., 2017 | Low | Controlling the age, sex. |

**S4.d:** Bias due to potential conflict of interest:

Bias due to potential conflict of interest resulting from support from a company, a study author or other entity with a financial interest were also assessed:

- Low: no conflict in the paper was mentioned
- Probably low: conflict of interest was not mentioned, but the laboratories that published the papers were affiliated with public research agencies or non-profit scientific institutions
- Probably high: there was insufficient information to allow a judgment of high risk of bias, but there was circumstantial evidence which suggests that the study was not free of support from a company, study author or another entity with a financial interest in the outcome of the study, as described by the criteria for a judgment of high risk of bias
- High: there was circumstantial evidence to suggest that the study was not exempt from support from a company, study author, or another entity with a financial interest in the outcome of the study

| **Bias** | **References** | **ladder** | **Information found in each article related to the bias** |
| --- | --- | --- | --- |
| Bias due to conflict of interest | Ackerman et al., 2019 | Low | Assistant Professor Ackerman was supported by a Public Health Early Career Fellowship from the National Health and Medical Research Council (NHMRC). The sponsor had no role in the study design, data collection, analysis, and interpretation, in the drafting of the manuscript, or in the decision to submit the manuscript for publication. There are no potential conflicts of interest regarding this work. |
|  | Dick et al., 2020 | Low | The authors have no conflicts of interest to declare. There are no external sources of support to report. It's a job for the US government. |
|  | Gelfman et al., 2009 | Probably low | Not mentioned- affiliated with public (research) agencies or scientific institutions that we judged to be free from commercial interests in the study findings. |
|  | Großschädl et al., 2014 | Low | The authors have declared that no competing interests exist. |
|  | Guido et al., 2020 | Low | The authors have declared that no competing interests exist. |
|  | Holte et al., 2003 | Probably low | Not mentioned- affiliated with public (research) agencies or scientific institutions that we judged to be free from commercial interests in the study findings. |
|  | Jimenez-Sanchez et al., 2010 | Probably low | Not mentioned- affiliated with public (research) agencies or scientific institutions that we judged to be free from commercial interests in the study findings. |
|  | Leijon et al., 2009 | Low | Competing interests: None declared. |
|  | Martin et al., 2014 | Probably low | Not mentioned- affiliated with public (research) agencies or scientific institutions that we judged to be free from commercial interests in the study findings. |
|  | Paloneva et al., 2015 | Low | The authors declare that they have no competing interests. |
|  | Pekkala et al., 2017 | Low | The authors declare that they have no competing interests. |
|  | Söderberg et al., 2018 | Low | Competing interests: None declared. |
|  | Solomon et al., 2007 | Low | This study had no competing financial interests. |
|  | Spitaels et al., 2020 | Low | The authors have declared no conflicts of interest. |
|  | Swain et al., 2020 | Low | The authors declare that there is no conflict of interest. |
|  | Yu et al., 2017 | Low | Competing interests: None declared. |

The potential presence of other biases was also assessed. We did not identify any other biases and rated all studies as probably low risk of other bias

**S4.e:** The quality of the statistical trend tests in the included articles was also assessed:

- Satisfactory Quality (SQ): trend or age-period-cohort (APC) model tests were performed.
- Probably Satisfactory Quality (PSQ): there was no mention of a trend test or APC, but confidence intervals of the incidence and / or prevalence of MSDs over time were reported.
- Probably Unsatisfactory Quality (PUQ): no mention of statistical tests and no confidence intervals reported.

| **Test quality** | **References** | **ladder** | **Information found in each article related to the quality of the test (tendency test and/or CI95%)** |
| --- | --- | --- | --- |
| Quality of the statistical trend tests | Ackerman et al., 2019 | PUQ | Not mentioned |
|  | Dick et al., 2020 | PSQ | Chi-squared test to statistically measure the temporal evolution of back pain and pain in the arm, but no consideration of age (table 2)- age is considered in Table 3 and 4 but without trend test. |
|  | Gelfman et al., 2009 | PSQ | CI95% for the incidence |
|  | Großschädl et al., 2014 | PUQ | Not mentioned |
|  | Guido et al., 2020 | SQ | The cohort models by period of age (APC) used to analyze and project the rates considering the processes on three scales of time, age and year of survey (period) and year of birth (cohort). |
|  | Holte et al., 2003 | PUQ | Not mentioned |
|  | Jimenez-Sanchez et al., 2010 | SQ | Statistical tests on trends (presence of p value) |
|  | Leijon et al., 2009 | PSQ | CI95% for the prevalence |
|  | Martin et al., 2014 | SQ | Statistical tests on trends (presence of p value) |
|  | Paloneva et al., 2015 | PUS | Not mentioned |
|  | Pekkala et al., 2017 | SQ | Statistical tests on trends (presence of p value) |
|  | Söderberg et al., 2018 | PUQ | Not mentioned |
|  | Solomon et al., 2007 | PUQ | Not mentioned |
|  | Spitaels et al., 2020 | SQ | The cohort models by period of age (APC) were used to analyze and project the rates considering the processes on three scales of time, age and year of survey (period), and year of birth (cohort). |
|  | Swain et al., 2020 | PUQ | Not mentioned |
|  | Yu et al., 2017 | PSQ | CI95% for the incidence |
